# The effects of ARBs, ACEIs and statins on clinical outcomes of COVID-19 infection among nursing home residents

**DOI:** 10.1101/2020.05.11.20096347

**Authors:** Anton De Spiegeleer, Antoon Bronselaer, James T Teo, Geert Byttebier, Guy De Tré, Luc Belmans, Richard Dobson, Evelien Wynendaele, Christophe Van De Wiele, Filip Vandaele, Diemer Van Dijck, Dan Bean, David Fedson, Bart De Spiegeleer

## Abstract

**Background.:** COVID-19 infection has limited preventive or therapeutic drug options at this stage. Some of common existing drugs like angiotensin-converting enzyme inhibitors (ACEi), angiotensin II receptor blockers (ARB) and the HMG-CoA reductase inhibitors (‘statins’) have been hypothesised to impact on disease severity. However, up till now, no studies investigating this association were conducted in the most vulnerable and affected population groups, i.e. older people residing in nursing homes. The purpose of this study has been to explore the association of ACEi/ARB and/or statins with clinical manifestations in COVID-19 infected older people residing in nursing homes.

**Methods and Findings.:** We undertook a retrospective multi-centre cohort study in two Belgian nursing homes that experienced similar COVID-19 outbreaks. COVID-19 diagnoses were based on clinical suspicion and/or viral presence using PCR of nasopharyngeal samples. A total of 154 COVID-19 positive subjects was identified. The outcomes were 1) serious COVID-19 defined as a long-stay hospital admission (length of stay ≥ 7 days) or death (at hospital or nursing home) within 14 days of disease onset, and 2) asymptomatic, i.e. no disease symptoms in the whole study-period while still being PCR diagnosed. Disease symptoms were defined as any COVID-19-related clinical symptom (e.g. coughing, dyspnoea, sore throat) or sign (low oxygen saturation and fever) for ≥ 2 days out of 3 consecutive days.

Logistic regression models with Firth corrections were applied on these 154 subjects to analyse the association between ACEi/ARB and/or statin use with the outcomes. Age, sex, functional status, diabetes and hypertension were used as covariates. Sensitivity analyses were conducted to evaluate the robustness of our statistical significant findings.

We found a statistically significant association between statin intake and the absence of symptoms during COVID-19 infection (unadjusted OR 2.91; CI 1.27-6.71; p=0.011), which remained statistically significant after adjusting for age, sex, functional status, diabetes mellitus and hypertension. The strength of this association was considerable and clinically important. Although the effects of statin intake on serious clinical outcome (long-stay hospitalisation or death) were in the same beneficial direction, these were not statistically significant (OR 0.75; CI 0.25-1.85; p=0.556). There was also no statistically significant association between ACEi/ARB and asymptomatic status (OR 1.52; CI 0.62-3.50; p=0.339) or serious clinical outcome (OR 0.79; CI 0.26-1.95; p=0.629).

**Conclusions.:** Our data indicate that statin intake in old, frail people could be associated with a considerable beneficial effect on COVID-19 related clinical symptoms. The role of statins and any interaction with renin-angiotensin system drugs need to be further explored in larger observational studies as well as randomised clinical trials.

## Introduction

Patients with serious and fatal COVID-19 infections are characterised by pneumonia-associated acute respiratory distress syndrome (ARDS) and multi-organ failure. The underlying mechanisms are linked to an imbalance between ACE and ACE2, as well as endothelial dysfunction (1-3) (**Figure 1**). Animal experiments have indicated that ARBs, ACEis or statins can prevent experimentally induced ARDS (4). These drugs are also likely to counteract the effects of sepsis-associated coagulopathy, elevated pro-inflammatory cytokines *(e.g*. IL-6) and sepsis-associated effects on pulmonary vascular permeability (5-12).

**Figure 1.**
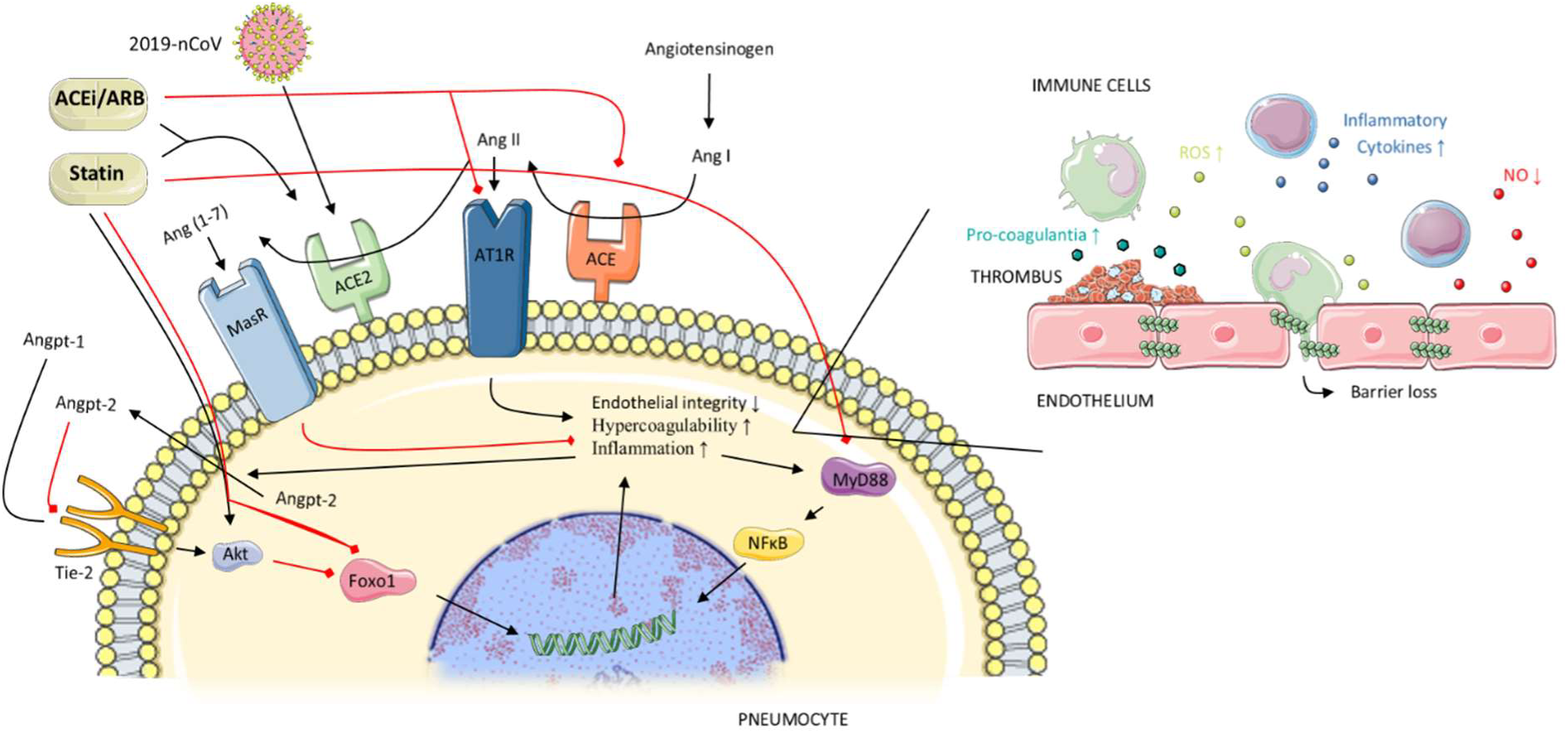
*Three mechanisms suggested for the effects of statins and ACEis/ARBs in preventing severe pulmonary disease in COVID-19. 1) Under normal conditions the Tie-2 receptor is continuously activated by Angiopoetin-1 (Angpt-1), which in turn activates Akt-kinase, leading to phosphorylation and hence inhibition of the transcription factor Foxo1. Unphosphorylated or active Foxo1 initiates the transcription of genes leading to increased inflammation, decreased endothelial barrier integrity and hypercoagulability. Angpt-2 is a partial antagonist of the Tie-2 receptor, stimulating inflammation, endothelial dysfunction and hypercoagulability. COVID-19 infection and ARDS are associated with increased Angpt-2 levels in blood, while statins simulate the Angpt-1 pathways. 2) The RAS system activates angiotensin-1 receptors (AT1R), stimulating inflammation, hypercoagulability and endothelial permeability. The Ang II-ACE2-Ang(1-7)-Mas receptor pathway counteracts the effects of this RAS system. COVID-19 enters the cell through ACE2 receptors, thereby decreasing these membrane-bound receptors, and relatively stimulating the RAS system. ACEis/ARBs inhibit the RAS system, while concomitantly increasing ACE-2 expression, which protects against ARDS. Statins also increase ACE-2 expression. 3) In ARDS there is an increase in the activation of the MyD88-NFkB inflammatory pathway. Statins preserve MyD88 at normal levels and down regulate NFkB. Black lines = stimulating effects; red lines = inhibiting effects.*

In a non-COVID-19 context, clinical investigators have observed that pneumonia patients who had been taking statins, ARBs or ACEis had improved survival (13, 14). Moreover, recent observational studies have reported similar findings for hospitalized COVID-19 patients (15-19). Recently, randomized controlled clinical trials have begun to evaluate the clinical effects of ARB, ACEi or statin treatment in hospitalized COVID-19 patients *(e.g*. NCT04348695, NCT04343001, NCT04351581). However, the estimated completion dates for these trials will be some time in 2021, and most will only consider ARB/ACEi monotherapy, *i.e*. not in combination with statins.

To our knowledge, no ARB/ACEi/statin studies have been or are being conducted among elderly nursing home residents, the most vulnerable individuals for COVID-19 morbidity and mortality. In Belgium, a country with a well-developed health care system, 3000 residents of nursing homes have died from COVID-19, with still around 100 residents a day currently dying (20). Estimates for the US suggest that almost 20% of all COVID-19 deaths have occurred in long-term care centers (21). Thus, every day without effective therapy comes at a high human cost.

We aimed to replicate the hospital findings to a frail, high-risk population living in nursing homes. While we wait for the results of prospective clinical trials, our findings allow us to make suggestions about the use of ACEis/ARBs and statins for these COVID-19 patients.

## Methods

This retrospective study conformed with all legal guidelines and the protocol was approved by the Ethical Committee of the Ghent University Hospital (reference BC-07671).

### Study design

The retrospective study cohort was defined as all (anonymised) residents at two elderly care homes with COVID-19 diagnosis based on clinical grounds and/or PCR lab testing from 1^st^ of March to 16^th^ of April. Both elderly care homes experienced COVID-19 outbreaks during this period. To determine the day of disease onset, structured and unstructured diagnostic records were analysed for symptoms suggesting COVID-19 infection. The first day of suggestive symptoms on two out of three consecutive days was considered as the day of disease onset. For the PCR-diagnosed residents, the suggestive symptoms used for disease onset were cough, shortness of breath (dyspnoea), sore throat, runny nose, general weakness, headache, confusion, muscle pain, arthralgia, diarrhoea, abdominal pain, vomiting, fever (T° > 37.6°C), increased oxygen need or low oxygen saturation (SpO2 ≤ 92%). In cases where no symptoms were mentioned (while still being PCR COVID-19-positive diagnosed), the date of nasopharyngeal sampling was used as the day of disease onset. For the clinically diagnosed residents without a confirmatory PCR lab test, the symptoms used for determining disease onset were defined more strictly, i.e., respiratory complaints (cough, shortness of breath, sore throat, runny nose), fever (T° > 37.6°C), increased oxygen-need or low oxygen saturation (SpO2 ≤ 92%).

The primary outcomes were 1) serious COVID-19, i.e. long-stay hospital admission (length of stay ≥ 7 days) or death (at nursing home or hospital) within 14 days of disease onset, and 2) asymptomatic, i.e. no disease symptoms as defined above throughout the whole study-period while still being PCR diagnosed.

All residents were stratified according to drug exposure to ACEi or ARB within 7 days before the day of disease onset or during the disease (prior to an outcome being reached). Specifically, we considered as treated all residents taking ≥ 2 days an ACEi (ramipril, lisinopril, enalapril, captopril, quinapril, imidapril, fosinopril, trandolapril) or ARB (candesartan, irbesartan, losartan, olmesartan, telmisartan, valsartan) up to 7 days before or 14 days after disease onset. An identical protocol was used to stratify according to drug exposure to statins (atorvastatin, fluvastatin, pravastatin, rosuvastatin, simvastatin).

We developed a mapping table based on clinical prescriptions to determine the diabetic and hypertension status of all residents. It was designed by a specialist in elderly care and validated by two independent physicians, one a general physician and the other a cardiologist.

The functional status of all residents was a dichotomous variable (high vs. low functioning). This definition was based on the available Katz scale for residents before day of disease onset. The Katz scale is a measure of independent activity of daily living.

### Data processing and quality control

Anonymized data were imported in a relational database for processing, using Extract, Transform, and Load (ETL) techniques. All received anonymized data were then evaluated on basic data quality attributes such as completeness (i.e., the extent of missing data) and accuracy (i.e., whether or not suspicious outliers were present in the individual attributes). Data were enriched with ATC codes for the included drugs. Suggestive symptoms were searched for, based on biometrical measurements as well as indications in text. For the later, basic Natural Language Processing (NLP) techniques were used. For the residents still in the hospital on the moment of data extraction, median imputation was used to estimate length of hospital duration. Two independent physicians manually verified all recorded symptoms as well as all data for a random subsample.

### Statistical Analysis

We calculated the distributions for dependent and independent variables for the total cohort using appropriate measures of central tendency and dispersion. For our main analysis, we investigated the association between ACEi/ARB and/or statin treatment and 1) serious disease, measured as long-stay hospital admission or death, or 2) asymptomatic disease using a series of logistic regressions applying Firth’s correction. This procedure has been used previously by our group and shown to be robust for low prevalence events and low-dimensional settings (16, 22, 23). We first explored the independent association between ACEi/ARB and both outcomes, as well as the association between statins and the same outcomes. Then we adjusted the models stepwise for age, sex, functional status, hypertension, and diabetes mellitus. All statistical analyses were performed using SAS 9.4 (SAS Institute, North Carolina, United States) and RStudio 3.5.2 (R Foundation for Statistical Computing, Vienna, Austria).

### Sensitivity Analyses

For the statistically significant associations, we also conducted sensitivity analyses to evaluate our modelling assumptions and the extent to which the population selection influenced the main results and conclusions. To control for the former (bias from modelling assumption) we conducted exact logistic regressions with models adjusted variable by variable. To control for the later (bias from population selection) we used the same modelling approach (logistic regression with Firth’s correction) to analyse only the PCR confirmed COVID-19+ residents.

## Results

The study cohort included 154 COVID-19-diagnosed residents aged 86±7 (mean±SD) years, evenly distributed over the two nursing homes (76 and 78 residents, respectively). Baseline characteristics are shown in **Table 1**. In our cohort (33% male), 20% were taking ACEis/ARBs (16% ACEi and 4% ARB), and 20% were taking a statin. Eight residents (5%) were taking both an ACEi/ARB and a statin. Important, none of the residents stopped ACEi/ARB or statin treatment on the day of disease onset and all continued taking their drugs during the follow-up period unless the clinical situation no longer allowed this. Also, none of the residents was taking other renin-angiotensin system (RAS)-associated drugs such as renin-inhibitors or neprylisine-inhibitors. Clinical symptoms detected by NLP in unstructured texts were all manually verified, with 22% false positives, mostly due to mentioned symptoms with more complex negations in the same sentence. Two physicians also independently evaluated manually all available data from a minimum of five random residents each. This resulted in no changes in the result-matrix.

**Table 1.** Characteristics of the study cohort. All variables are shown as N (% of column), except age which is mean in years (SD). ACEi = Angiotensin converting enzyme inhibitor; ARB = Angiotensin receptor blocker.

| Sample characteristics | Total (N = 154) | ACEi/ARB (N = 30) | No ACEi/ARB (N = 124) | Statin (N = 31) | No statin (N = 123) | Symptoms (N = 113) | No symptoms (N = 41) | Serious COVID (N = 37) | Non-serious COVID (N = 117) |
| --- | --- | --- | --- | --- | --- | --- | --- | --- | --- |
| Age | 85.9 (7.2) | 86.2 (6.6) | 85.8 (7.4) | 85.6 (5.3) | 85.9 (7.6) | 86.0 (7.4) | 85.6 (6.6) | 86.8 (6.8) | 85.6 (7.3) |
| Male | 51 (33.1%) | 12 (40.0%) | 39 (31.5%) | 10 (32.3%) | 41 (33.3%) | 41 (36.3%) | 10 (24.4%) | 12 (32.4%) | 39 (33.3%) |
| On ACEi/ARB | 30 (19.5%) | 30 (100%) | 0 (0%) | 8 (25.8%) | 22 (17.9%) | 20 (17.7%) | 10 (24.4%) | 6 (16.2%) | 18 (20.5%) |
| On statin | 31 (20.1%) | 8 (26.7%) | 23 (18.5%) | 31 (100%) | 0 (0%) | 18 (15.9%) | 14 (34.1%) | 6 (16.2%) | 25 (21.4%) |
| Low functioning | 137 (89.0%) | 23 (76.7%) | 114 (91.9%) | 26 (83.9%) | 111 (90.2%) | 106 (93.8%) | 31 (75.6%) | 35 (94.6%) | 102 (87.2%) |
| Diabetes mellitus | 28 (18.2%) | 6 (20.0%) | 22 (17.8%) | 10 (32.3%) | 18 (14.6%) | 18 (15.9%) | 10 (24.4%) | 7 (18.9%) | 21 (17.9%) |
| Hypertension | 39 (25.3%) | 28 (93.3%) | 11 (8.87%) | 8 (25.8%) | 31 (25.2%) | 29 (25.7%) | 10 (24.4%) | 10 (27.0%) | 29 (24.8%) |
| Symptoms | 113 (73.4%) | 20 (66.7%) | 93 (75.0%) | 17 (54.8%) | 96 (78.0%) | 113 (100%) | 0 (0%) | 36 (97.3%) | 77 (65.8%) |
| Serious COVID | 37 (24.0%) | 6 (20.0%) | 31 (25.0%) | 6 (19.4%) | 31 (25.2%) | 36 (31.9%) | 1 (2.44%) | 37 (100%) | 0 (0%) |

Of the 154 residents, 41 remained asymptomatic during the study period, i.e. 27% of the total cohort and 47% of the PCR-tested COVID-19+ residents. These numbers are similar to those from another study in a similar population (24). Thirty-seven residents (24%) experienced serious COVID-19. Although this serious outcome number seems high compared to other outpatient population studies, in view of the very vulnerable population this is not surprising (25, 26). Among residents treated with ACEis or ARBs, 10/30 (33%) remained asymptomatic vs. 31/124 (25%) of those without such treatment. Residents taking statins remained asymptomatic in 45% of the cases (14/31) vs. 22% (27/123) of those not taking statins. Evaluating COVID-19 severity, 20% (6/30) of the residents treated with ACEi/ARB died or were admitted to hospital for long-stay vs. 25% (31/124) of those without such treatment. Residents taking statins experienced serious COVID-19 in 19% of the cases (6/31) vs. 25% (31/123) of those not taking statins. Interestingly, six of eight residents (75%) taking the ACEi/ARB and statin combination remained asymptomatic throughout the study period. Only one of them (13%) experienced serious COVID-19.

Although not reaching statistical significance, findings from unadjusted logistic regression suggested a potential beneficial effect on COVID-19 symptoms among residents taking ACEis or ARBs (OR 1.52; CI 0.62-3.50; p=0.329). Odds ratios adjusted for age, sex, functional status, diabetes and hypertension were of similar magnitude (**Table 2**). The results for the statins were most interesting, as we observed a clear and statistically significant association between statin intake and asymptomatic status (unadjusted OR 2.91; CI 1.27-6.71; p=0.011). This association was partially attenuated but remained statistically significant when adjusted for gender, age, functional status, diabetes and hypertension (**Table 2**).

**Table 2.** Summary of odds ratios for the asymptomatic COVID-19 infection using logistic regression with Firth’s correction.

| Drug treatment | Adjustments | OR (95% CI) on drug vs. no drug | P-value |
| --- | --- | --- | --- |
| ACEi/ARB | - | 1.52 (0.62-3.50) | 0.339 |
|  | Age, sex | 1.61 (0.65-3.80) | 0.283 |
|  | Age, sex, functional status | 1.35 (0.51-3.31) | 0.521 |
|  | Age, sex, functional status, diabetes mellitus, hypertension | 2.72 (0.59-25.1) | 0.242 |
| Statins | - | 2.91 (1.27-6.71) | 0.011 |
|  | Age, sex | 2.88 (1.26-6.83) | 0.013 |
|  | Age, sex, functional status | 2.87 (1.23-7.07) | 0.016 |
|  | Age, sex, functional status, diabetes mellitus, hypertension | 2.65 (1.13-6.68) | 0.028 |

We also examined associations between ACEis/ARBs and statins, and serious COVID-19. Although the available data failed to reach statistical significance, the directionality of the odds ratios suggested a potential beneficial clinical effect of both ACEi/ARB and statins on serious COVID-19 outcome. All odds ratios (unadjusted as well as adjusted for covariates), were between 0.48 (CI 0.10-1.97; p=0.316) and 0.84 (CI 0.27-2.14; p=0.736) (**Table 3**).

**Table 3.** Summary of odds ratios for the serious COVID-19 infection using logistic regression with Firth’s correction.

| Drug treatment | Adjustments | OR (95% CI) on drug vs. not drug | P-value |
| --- | --- | --- | --- |
| ACEi/ARB | - | 0.79 (0.26-1.95) | 0.629 |
|  | Age, sex | 0.78 (0.25-1.93) | 0.610 |
|  | Age, sex, functional status | 0.84 (0.27-2.14) | 0.736 |
|  | Age, sex, functional status, diabetes mellitus, hypertension | 0.48 (0.10-1.97) | 0.316 |
| Statins | - | 0.75 (0.25-1.85) | 0.556 |
|  | Age, sex | 0.75 (0.25-1.86) | 0.564 |
|  | Age, sex, functional status | 0.77 (0.25-1.91) | 0.597 |
|  | Age, sex, functional status, diabetes mellitus, hypertension | 0.75 (0.24-1.87) | 0.559 |

We did not undertake regression analyses on the combined ACEi/ARB+statin group as there were only eight residents in our cohort; nor did we undertake separate analyses for the ACEi or ARB groups; only six residents were treated with an ARB.

Sensitivity analyses were conducted on the statistically significant association between statins and symptoms. We found that estimates of the impact of statin treatment on asymptomatic status were consistently of the same magnitude and statistically significant as the original analyses.

## Discussion

There are currently no licensed antiviral treatments for COVID-19 approved. Also the development of COVID-19 vaccines will take time. Moreover, there is no information on when sufficient vaccine supplies will become widely available. Recently, the World Health Organization (WHO) communicated a Solidarity “megatrial” evaluating four broad-spectrum antiviral agents. Among them, the broad-spectrum experimental antiviral drug remdesivir was shown to have low efficacy against Ebola and dropped from further study, although a recent report of its compassionate use in serious COVID-19 was favourable (27). Lopinavir and ritonavir (a protease inhibitor combination used to treat HIV patients) were ineffective in a Chinese clinical trial (28). A lot of attention has gone to chloroquine and hydroxychloroquine. Unfortunately prospects for their success against COVID-19 are not good (29). Convalescent sera, obtained from recovered COVID-19 patients, might be an option to treat acute COVID-19 infections (30), but its implementation will be cumbersome and unlikely to become widely available. The first clinical trials of ACEi/ARB and statin treatments in hospital settings have been initiated within the past month. While we await the results of these trials, which are expected in 2021, this retrospective study should be regarded as both timely and complementary, as it has focused on a frail, non-hospitalised population and demonstrated clinical findings on the use of ACEi/ARB/statins using real world data.

Although statistically not significant, overall both ACEi/ARB and statins show clinical beneficial odds ratios for the outcome serious COVID-19 in elderly people who live in nursing homes. The results for statins and symptoms are most convincing, i.e. large effect sizes which are statistically significant. Statins are most frequently used to prevent cardiovascular diseases. The safety profile of statins is well known and excellent, even in the old population. Moreover, these drugs are relatively inexpensive and widespread, some even as food supplements as red yeast rice, making them easily available throughout the world. Although this observational study does not have the power of a randomized controlled clinical trial, in the current absence of other valuable therapies and considering the benefit-risk balance, an older person living in a nursing home could consider taking a statin if at high COVID-19 infection risk. Currently, therapeutic decisions for COVID-19 patients are driven by observational studies (31, 32). In any case, based on our results, we recommend against stopping statins in patients who are COVID-19-infected.

The combination of ACEi/ARB and statin treatment seemed to have additive beneficial effects on symptoms and serious disease outcome: six of eight residents taking the combination remained asymptomatic and only one of them developed serious COVID-19. Although this result is promising, our sample size was too small to allow us to draw firm conclusions.

One strength of this study is the specific population, i.e., old people (mean age > 85years) residing in nursing homes. Although they are considered highly vulnerable to COVID-19 clinical outcomes, no study has yet reported on the effects of ARB/ACEi and/or statin treatment on COVID-19 in this population. Extracting reliable data from nursing homes with COVID-19 outbreaks is far more cumbersome than extracting data from hospitals. Another strength is that drug treatment was based on real intake, in contrast to most hospital-based studies that use prescriptions as proxies for drug treatment. Lastly, in contrast to most hospital studies, asymptomatic COVID-19 patients were included in the study. People admitted to hospitals are evidently always symptomatic.

One limitation of our study is its relatively small cohort size. Consequently, absence of statistical significance should be interpreted with caution. However, the consistency in the observed effect sizes, even without statistical significance due to small sample size, should be considered in the overall evaluation. As number of cases increase, further analyses will be undertaken to better understand our findings and confirm these associations. Also, another limitation was the lack of other potential confounders, including chronic kidney injury and BMI. Finally, our results apply to a very specific population (elderly people living in nursing homes) and cannot be generalized to other groups such as young people or hospitalized people.

## Conclusions

Our study, based on available data, indicates that in elderly nursing home residents, statin treatment is associated with beneficial effects on COVID-19-related clinical symptoms. Although not statistically significant, our findings also suggested that statin treatment in combination with an ACEi or ARB was associated with less severe clinical outcomes. In the light of these findings, a prudent recommendation is to continue or initiate statin treatment for older people residing in nursing homes and at high risk for COVID-19 infection.

## Data Availability

available from corresponding author

## Acknowledgements

We thank all of the staff of VZW Zorg-Saam Zusters Kindsheid Jesu for their daily care of older people and for their collaboration on this study during these difficult times. We also thank the staff of the Corilus Health IT Center who helped with the data extraction.

## Notes

### Competing Interest Statement

The authors have declared no competing interest.

### Funding Statement

ADS is supported by a grant of Research Foundation Flanders (FWO) (grant number 1158818N).

